# A double-blind, crossover, non-inferiority randomized controlled trial where primary care providers and patients compare human- and AI-generated digital health messages: the AI-CARE study protocol

**DOI:** 10.64898/2025.12.19.25341381

**Authors:** Audrée Lemieux, Stephen A Kutcher, Borris Rosnay Galani Tietcheu, Gretchen Seitz, Jason Trickovic, Douglas Archibald, Sylvie Grosjean, William Hogg, Sharon Johnston

## Abstract

**Introduction:** Primary care is facing multiple crises, including an increase in health misinformation. Digital health messaging by primary care providers has been shown to reach a diverse patient population. With the uptake of Generative Artificial Intelligence (GenAI) usage in healthcare, there is an important opportunity to rapidly create messages that are tailored to different populations and conditions. However, thoroughly assessing AI-generated content is essential, as GenAI raises concerns regarding its accuracy, understandability, actionability, and bias perpetuation. We aim to investigate whether digital health messages created by GenAI are evaluated as noninferior compared to those created by human experts.

**Methods and analysis:** The AI-CARE (AI to Create Accessible and Reliable patient Education materials) study is a double-blind, crossover, non-inferiority randomized controlled trial. Data collection began on May 30, 2025, and is expected to be completed at the end of April 2026. Over 12 months, 192 messages on 48 topics will be written: half by primary care and public health experts and half by a GenAI tool (OpenAI’s ChatGPT). Review Panels composed of 24 primary care providers and 24 patients will evaluate these messages using an Evaluation Grid developed to assess the messages’ quality of information, adaptation to the target audience, relevance and usefulness, and readiness to be shared with patients. Evaluations will be completed via online REDCap surveys and the order in which the 192 messages appear will be randomized and will vary between individuals. Participants and analysts will be blinded to the generation source. The primary outcome will be the Clarity and Understandability score.

**Ethics and dissemination:** The Research Ethics Boards of the Hôpital Montfort (24-25-11-038) and the University of Ottawa (S-12-24-11153) formally approved this study in December 2024. Reported data will be grouped and anonymized for dissemination in peer-reviewed scientific journals and conferences.

**Trial registration number:** NCT06997107

**ARTICLE SUMMARY:** *Strengths and limitations of this study:*

- The AI-CARE study allows for within-participant comparison between human- and AI-generated digital health messages, minimizing variability due to individual differences.
- The Review Panels are diverse and composed of primary care providers and patients currently practicing in or using the healthcare system in five Canadian provinces.
- The developed Evaluation Grid allows for the assessment of multiple aspects related to digital health messages: quality of information, adaptability to the target audience, relevance and usefulness, and readiness to be shared with patients.
- One limitation is that messages generated by AI are created using only one LLM (Open AI’s ChatGPT).
- Due to the nature and location of recruitment, we may introduce selection bias (participants already engaged in research and interested in digital communication and AI) and the racial diversity of our study population may be limited.

## INTRODUCTION

As the initial point of contact, primary care providers play a trusted role in providing accurate information about emerging health topics [1,2]. Yet, few practices have the time, digital communications expertise, or resources to routinely develop or share proactive health information with their patients that addresses the wide range of concerns for which people seek information. Moreover, primary care is facing a confluence of crises, including shortages in health human resources, increasing patient complexity, and rampant health misinformation [3]. Enabling primary care practices to rapidly provide timely, accurate, and accessible digital health messages that are tailored to different populations and conditions can help improve access to health information, empower the patient-provider relationship [4], and combat the epidemic of health misinformation [5–7].

In the past five years, the Canadian Primary Care Information Network (CPIN) (see https://cpin-rcip.com/), a bilingual research program and infrastructure that develops and studies approaches for digital patient engagement in primary care [8], has sent digital health messages to over 90,000 patients across Canada [9]. Topics for patient education materials included Lyme disease prevention, physical activity, mental health, or updates on COVID-19 and flu vaccines. Recent data showed that this digital patient engagement can reach a diverse population, including individuals typically hard to reach, such as older adults, members of racialized communities, people in rural areas, or individuals with multiple medical conditions [9]. As more practices adopt secure patient portals, email communication with patients, digital visit summaries, or text messaging for appointment or health reminders [10–15], determining what health information patients seek and the most effective ways to communicate it to them is essential to ensure this growing technology benefits both patients and primary care teams.

Since the release of OpenAI’s ChatGPT (Chat Generative Pre-trained Transformers) in November 2022 [16], Generative Artificial Intelligence (GenAI), and in particular large language models (LLMs), have rapidly gained attention for their ability to predict and, in turn, create human-like content mimicking aspects of cognition, such as reasoning [17–19]. Because of these abilities, they are increasingly becoming integrated in primary care and have the potential to offer more personalised health care [20–23]. For instance, GenAI has been used to help draft patient inbox messages [24], reason through test results and differential diagnoses [25,26], and generate medical notes using AI Scribe tools [27–29].

In patient education, the development of medical GenAI-powered chatbots has been one of the major focuses to provide general health information [6,19,30,31]. While Med-PaLM-2, Me-LLaMa, and other medically trained LLMs have been shown to produce information with higher accuracy due to the quality of their training data [32–34], readily accessible LLMs such as ChatGPT or Gemini are trained on both reliable and possibly unreliable online information [31]. This raises concerns about their potential for propagating health misinformation, especially in the context of patient care. GenAI is also known for occasionally creating false information due to its data quality and probabilistic architecture, presenting generated content in a factual and convincing way (termed “AI hallucinations”) [19–21,31]. Prompt strategies can help mitigate these hallucinations [35]. Key communication concepts, such as understandability, appropriate use of appeal and tone, and actionability, also need to be considered if these emerging tools are to extend the reach of health care providers’ communication [36–39]. Additionally, research on GenAI as a health communication tool must vigilantly prevent the amplification of bias and the worsening of health inequities [40–42].

Many experts emphasize that AI should complement rather than replace the clinical expertise of real, trained providers [43–46]. Several studies have implemented safety mechanisms, such as requiring mandatory approval from a provider before sending any AI-generated content to patients [47]. This human-in-the-loop mechanism reflects a growing consensus that human oversight with AI-supported communication remains essential.

Given these concerns, several studies have explored whether AI-generated content can meet acceptable standards of safety and quality. For the most part, these studies focused on only one or two key elements, such as readability, understandability, and/or accuracy, and only a few explored actionability, a key element of effective health communication [48–55]. Results indicated that messages to patients or patient education materials generated by AI and humans were comparable for readability, clarity, and quality [49,53,56,57], although trust in digital medical advice decreased once people knew it was generated by AI [56,57]. Among these studies, only one included primary care providers [49]. Very few studies to date investigating AI communication tools in healthcare involve patients, the actual intended audience for general health information [50,58].

As part of the CPIN research program, we are conducting the AI to Create Accessible and Reliable patient Education materials (AI-CARE) randomized controlled trial, where both primary care providers and patients evaluate human- and AI-generated digital health messages on four main aspects: quality of information, adaptation to the target audience, relevance and usefulness, and readiness to be shared with patients. We aim to investigate whether the understandability of AI-generated digital health messages is noninferior compared with those created by human experts. The results will provide insights into how GenAI can be routinely implemented in primary care to support digital health message creation and patient education.

## METHODS AND ANALYSIS

### Trial design

The AI-CARE study is a double-blind, crossover, non-inferiority randomized controlled trial. Figure 1 describes the overall methodology for each of the 12 months. The AI-CARE Review Panels composed of primary care providers and patients (now referred to as “panelists”) will evaluate 192 messages on 48 different topics (16 messages per month on 4 different topics). For each of the 48 topics, both a human expert (“human-generated”) and a GenAI tool (now simply referred to as “AI”; “AI-generated”) will produce a short-format message (<900 characters) and a longer-format message (∼1 page), for a total of 192 messages. This design allows panelists to directly compare human- and AI-generated content across formats most commonly used in digital primary care communication [59,60]. This study will be conducted in Canada’s two official languages, English and French.

**Figure 1.**
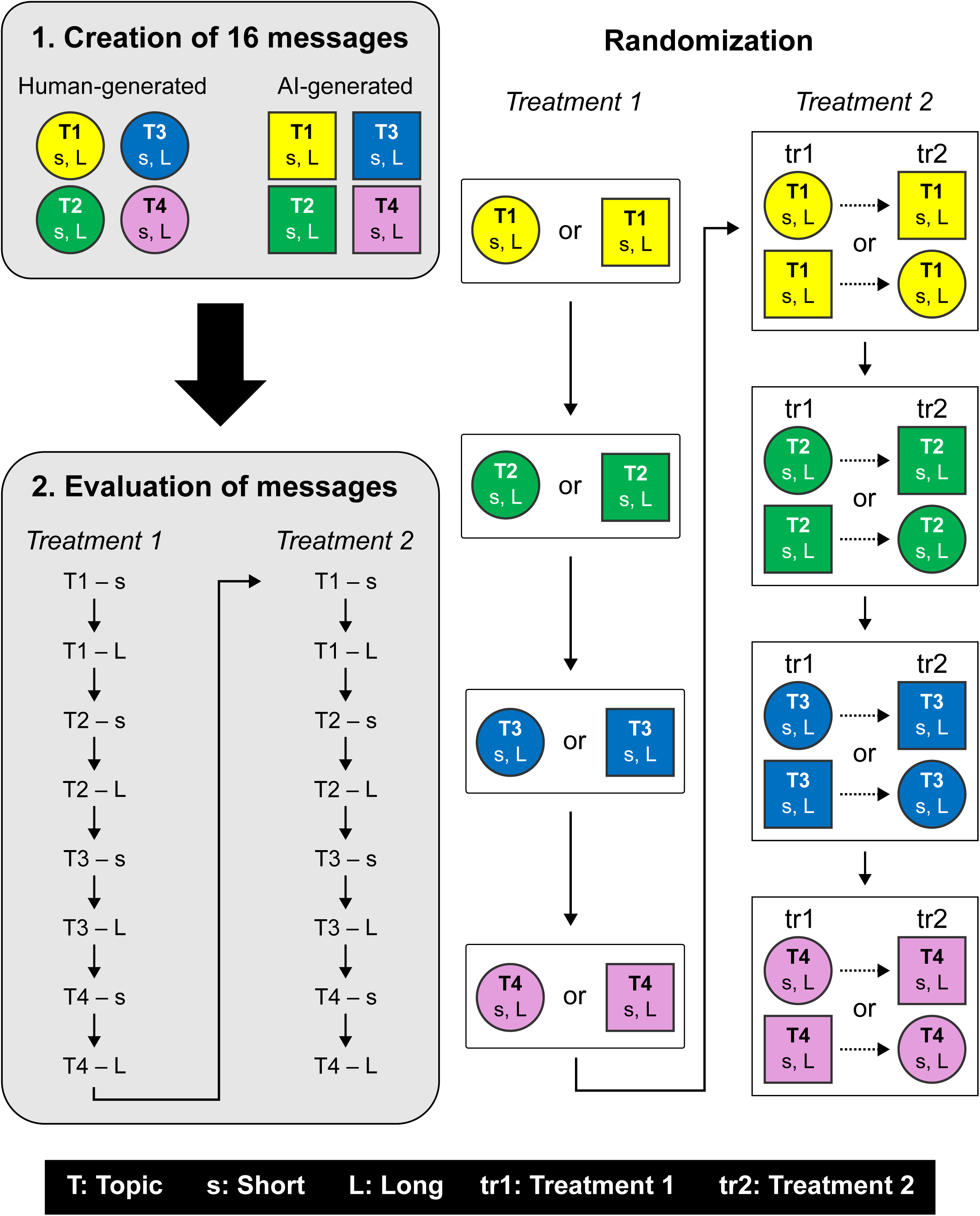
Overall monthly AI-CARE methodology. Each month, (step #1) 16 digital health messages will be generated by humans and AI, and (step #2) will be subsequently evaluated by Review Panels composed of primary care providers and patients. Review Panels will evaluate all 16 messages, but the order in which messages appear will be randomized and will vary between individuals. The generation source (human or AI) is first randomly assigned for Topics 1 to 4 (“Treatment 1”), and is later followed by the short and long messages from the alternate source (“Treatment 2”). Randomization is done at the topic level and for each individual. T: Topic; s: Short; L: Long; tr1: Treatment 1; tr2: Treatment 2.

### Data collection at baseline

Figure 2 describes the participant timelines. All panelists’ sociodemographic information will be collected at onboarding (see online supplemental material 1 for survey questions). Specifically, we will collect age, gender, sexual orientation, racial or cultural group, the first three characters of postal code, the language most spoken at home, and the official language of Canada in which allophones (i.e., most spoken language at home is neither English or French) are most comfortable receiving their healthcare services in. In addition, patient panelists will be asked about their highest education level attained, gross annual income, and status of caregiver. For primary care provider panelists, the profession, first three characters of the workplace postal code, and the number of years of practice will also be collected. Furthermore, at baseline, primary care providers will be asked about their usage of GenAI in their general practice and/or to create or adapt patient education materials (see online supplemental material 1 for survey questions).

**Figure 2.**
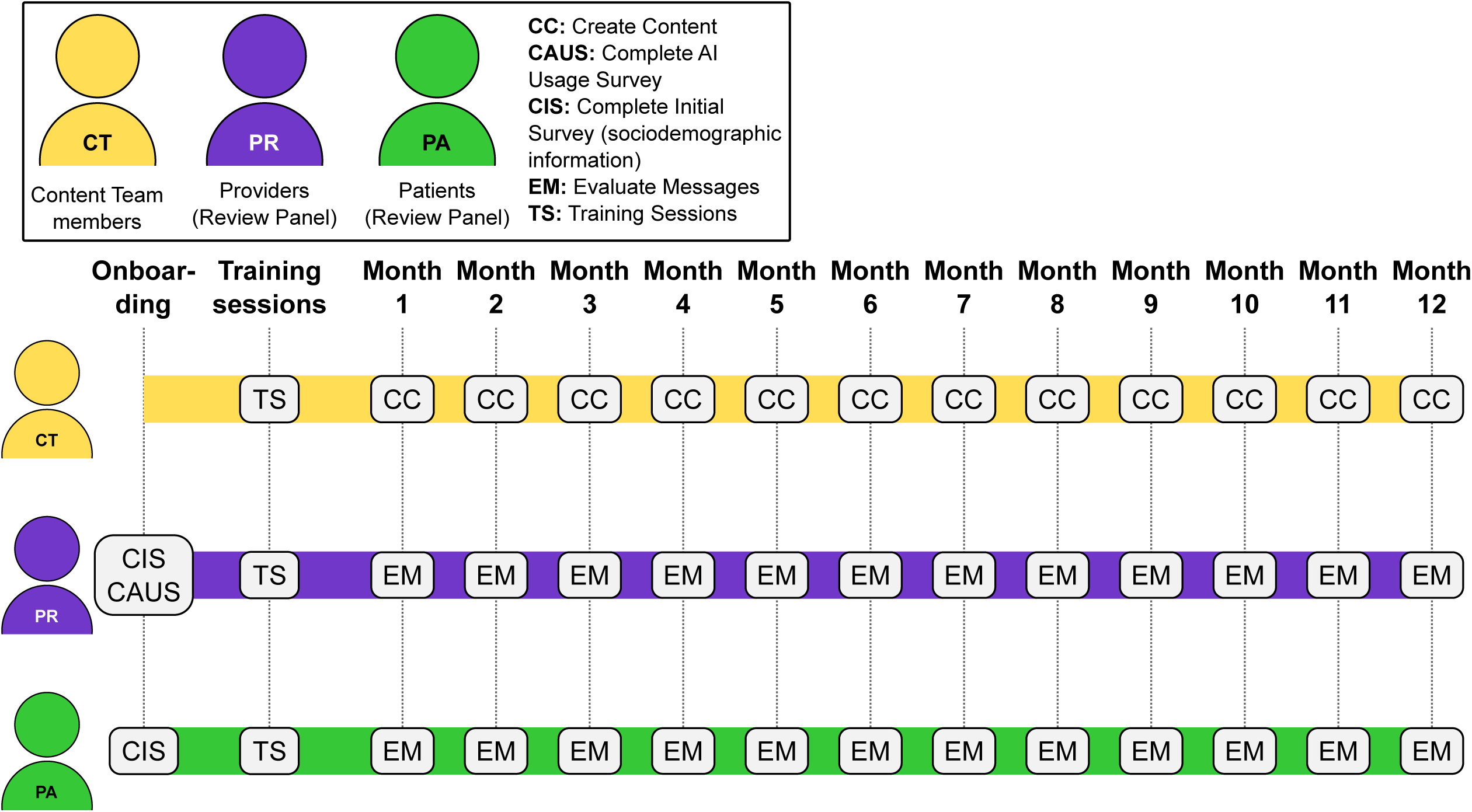
Participant timeline. This timeline illustrates the schedule for a typical participant. Individuals who join after data collection has begun may follow a condensed timeline to complete all evaluations. CC: Create Content; CAUS: Complete AI Usage Survey; CIS: Complete Initial Survey (sociodemographic information); EM: Evaluate Messages; TS: Training Sessions.

### Creation of digital health messages

“Human” digital health messages will be generated by experts specializing in primary care, public health, and/or health communication (now referred to as the “Content Team”). Each month, alternating Content Team members will write messages on one of four different topics aligned with their expertise (Figure 2), in two formats: a short, text-messaging format (900 characters or less), and a one-page handout for patients. Suggestions for topics have been provided by participating CPIN primary care providers and patients, and topics will be determined based on the Content Team member specialization. Messages for a given topic will be written in the Content Team members’ mother tongue or preferred language (English or French) and then translated to the other language (French or English) by a professional health communication translator. Prior to the start of content development, the Content Team received a training session on how to write patient education materials by a communications expert, who is responsible for creating or reviewing nearly all CPIN digital health messages [9]. The Content Team members will be prohibited from using any GenAI tools in crafting their messages. Grammar or spelling checkers will be allowed, as long as any GenAI features are disabled by the user. Each Content Team member will also specify the amount of time it took to write the messages.

Simultaneously, “AI” messages will be generated by a GenAI tool, which will be given the same instructions, prompts, and topics as the Content Team. Periodic updates on instructions will be provided to Content Team members and the GenAI tool. Two sets of prompts and instructions (one in English and one in French) have been developed. For each topic, the language of prompts will correspond to the language in which the Content Team member wrote their original message. The messages generated will then be professionally translated into the other language, as previously described. The GenAI tool will also be asked for the amount of time it took to craft the messages.

As this is a rapidly evolving field, we sought a GenAI tool that performed well in early evaluations of health-related queries in 2024-2025 and that would also be available to a typical primary care provider. We opted for OpenAI’s ChatGPT [61], as evidence before the beginning of content development showed that it regularly outperformed other LLMs such as Gemini [62], Google Bard, Microsoft Bing, and Llama [63–66] in health content creation. Given the rapid updates of these tools and that previous versions are not always available, we decided to systematically use the latest version of the GenAI tool each month (GPT-4o for months 1 to 3, GPT-5 for months 4 to 6, GPT-5.1 starting month 7).

We created a series of seven standardized prompts for the GenAI tool, which provide detailed, step-by-step instructions for health message generation that can be easily reproduced and integrated into routine primary care. The prompts were developed based on the most recent best practices for prompt engineering, such as using an iterative approach, giving context to clearly explain the project and its objectives, providing clear and specific instructions, asking the GenAI tool to assume a profession or qualifications of the simulated user (“primary care provider” in this context), and requesting the GenAI to improve its response based on its previous output [67–70]. All messages will be generated using the same set of prompts, with only the specific requirements (i.e., topic, target audience, time of the year) varying. To refresh the context every time and avoid the GenAI to get mixed up between topics, each of the 48 topics will be generated in a separate ChatGPT thread. The seven prompts can be found in Table 1.

**Table 1.** Prompts given to the GenAI tool and their objectives. [TBD] designate the message’s specific requirements that change each month.

| # | Prompt | Objectives of the prompt |
| --- | --- | --- |
| 1 | Hi ChatGPT! As a family doctor, I'm collaborating with the Canadian Primary Care Information Network research team on the innovative AI-CARE project. Together, we're exploring how artificial intelligence can help create powerful, personalized health promotion messages for patients—messages that are as effective as those written by real family doctors. I am seeking your assistance in drafting the best health promotion messages that will be sent to all my patients. Included are two AI-CARE training resources designed to guide the creation of impactful digital communications in primary care. Please let me know if you clearly understand the instructions. | Introduce the AI-CARE project and our objectives. |
| 2 | Wonderful! Now, imagine you are an expert in health communication and I, a licensed family doctor. Could you please help me with this while following the specific requirements given below?<br>1. The message must include a short and catchy title.<br>2. It should start with “Dear Patient” and end with a concluding sentence calling to action and ending with “Your Family Doctor”.<br>3. The message must be a maximum of 850-900 characters, spaces included.<br>4. The message will be sent to all [TBD]<br>5. The topic will be on: [TBD]<br>6. The time for the message is: [TBD]<br>7. The message should be able to be read by a 12-year-old reading level.<br>8. Where appropriate, include links to reliable sources for more information (use one link in short messages, and 1–3 links (or, exceptionally, more) for longer messages if suitable)<br>9. The link must include an existing full URL (beginning with https:// or http://) and should be a reliable Canadian resource. The link can be found on the websites of provincial or federal government agencies, as well as reputable medical schools, hospitals, and professional health organizations, such as the Canadian Medical Association, the Canadian Paediatric Society, CHEO, Unity Health Toronto, the University Health Network, and the Centre for Addiction and Mental Health. Other organizations, like CPIN and JAMA, may also provide useful links.<br>10. The message should contain only factual and verified | Create a first short message given a list of requirements. |
|  | information. Please do not use social media platforms as a source of material, especially not Facebook, Reddit, or Twitter.<br>11. Please use a tone and message appeal appropriate for this target audience<br>12. Provide the time spent to generate that message and not the human-like estimated duration.<br>13. Avoid using em dashes; whenever possible, use commas, brackets or parentheses instead. |  |
| 3 | Could you please create three different versions of the message containing different links, review all of them based on the guidelines provided to determine which one is the best, and finally, extract the best parts to create a new message, keeping in mind the previous instructions given? | Create three versions of the message and create a new, improved one. |
| 4 | Can you improve that message and compare both versions? | Create a new, improved short message. The output of prompt #4 is uploaded in the REDCap surveys for short messages. |
| 5 | Based on the improved version, could you please adapt the content for another target audience, like “Teenagers” or “Immunocompromised people”? | For more information only. |
| 6 | Could you please generate the long version of the improved message, following the previous specifications? Please remember to provide the specific time spent creating the message, as specified above. | Create the long version of the message. The output of prompt #6 is uploaded in the REDCap surveys for long messages. |
| 7 | What makes this your best long message? | For more information only. |

When applicable, small typos, grammatical errors, and non-functioning hyperlinks will be corrected in all messages. Of note, at the beginning of the data collection, the use of em dashes was becoming increasingly discussed in popular media to be a “signature” of ChatGPT and other LLMs [71,72]. Starting in month 5 of the study, we introduced a new requirement in prompt 2 to instruct both the Content Team and the GenAI tool to avoid using em dashes and prioritize the use of commas, brackets, and parentheses instead. This was done to avoid participants predicting which messages are generated by humans or AI, which could bias their evaluations [56,57].

### Evaluation of digital health messages by provider and patient Review Panels

To evaluate the generated content, two Review Panels (one of primary care providers and one of primary care patients) will assess the human- and AI-generated messages (Figure 2). Panelists will be blinded to the message generation source (“human” or “AI”), as described in the “Randomization and Blinding” section.

Short messages will always be shown before longer messages to minimize potential bias from exposure to more detailed content and to ensure that messages are effectively assessed given their length (Figure 1). For each topic, panelists will receive the short and long messages from a randomly assigned generation source first (“Treatment 1;” Human [H] or AI [A]), later followed by the short and long messages from the alternate source (“Treatment 2”). Panelists will review all topics under Treatment 1 in a sequential order (e.g., Topic 1 [H] --+ Topic 2 [A] --+ Topic 3 [A] --+ Topic 4 [H]), followed by the same topics under Treatment 2 in the same order (e.g., Topic 1 [A] --+Topic 2 [H] --+ Topic 3 [H]--+ Topic 4 [A]). This allows “washout periods” to be introduced between evaluations for each topic.

### The Evaluation Grid (outcome measurement tool)

Table 2 describes the 24 (primary care providers) and 19 (patients) statements presented in the Evaluation Grid, the outcome measurement tool used by panelists to assess messages. Based on the QUEST [73] and CLEAR [70] frameworks and adapted from the work of digital health communication experts [33,74–79], the Evaluation Grid was designed to evaluate the message content’s adaptability to its target audience (statements #6 to #19) [74–79], its relevance and usefulness to patients (statements #20 to #22) [74], and whether the message is ready to be shared with patients in its current form (statement #25). Additionally, primary care providers will be asked to evaluate the accuracy, reliability, and completeness of the information provided (statement #1 to #5) [33], as well as whether the sending of this message would increase administrative burden (e.g., higher number of calls to request appointments) on the practice (statement #24). Patients will not be asked to evaluate the quality of information and will simply assess whether they believe they have detected any inaccuracies (statement #23). For statements #1 to #24, the Evaluation Grid uses a Likert scale from 1 to 4 (1: “Strongly Disagree”; 2: “Disagree”; 3: “Agree”; and 4: “Strongly Agree”) and an additional option for “Not Applicable.” The Evaluation Grid was further pre-validated with panelists during training sessions.

**Table 2.**
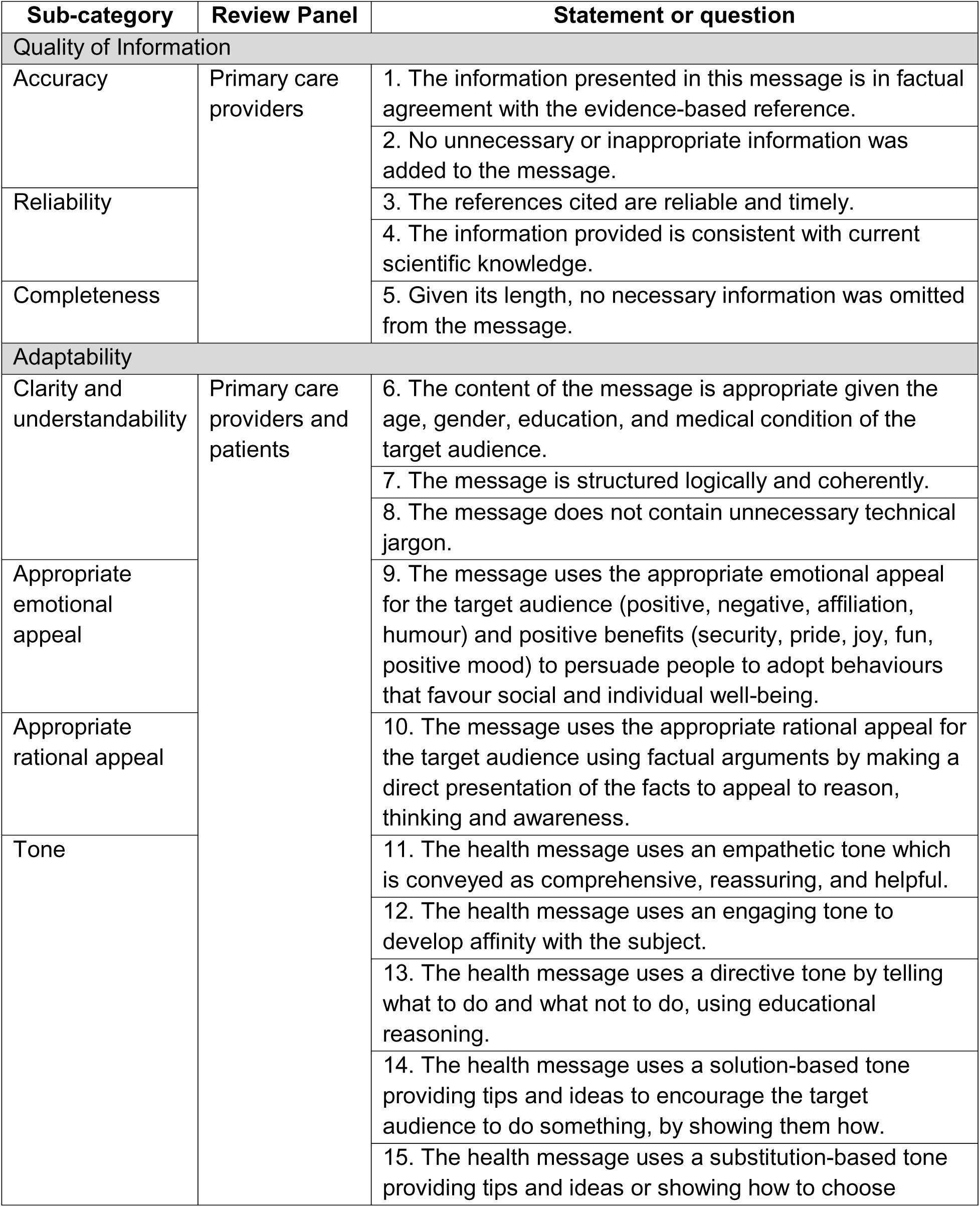
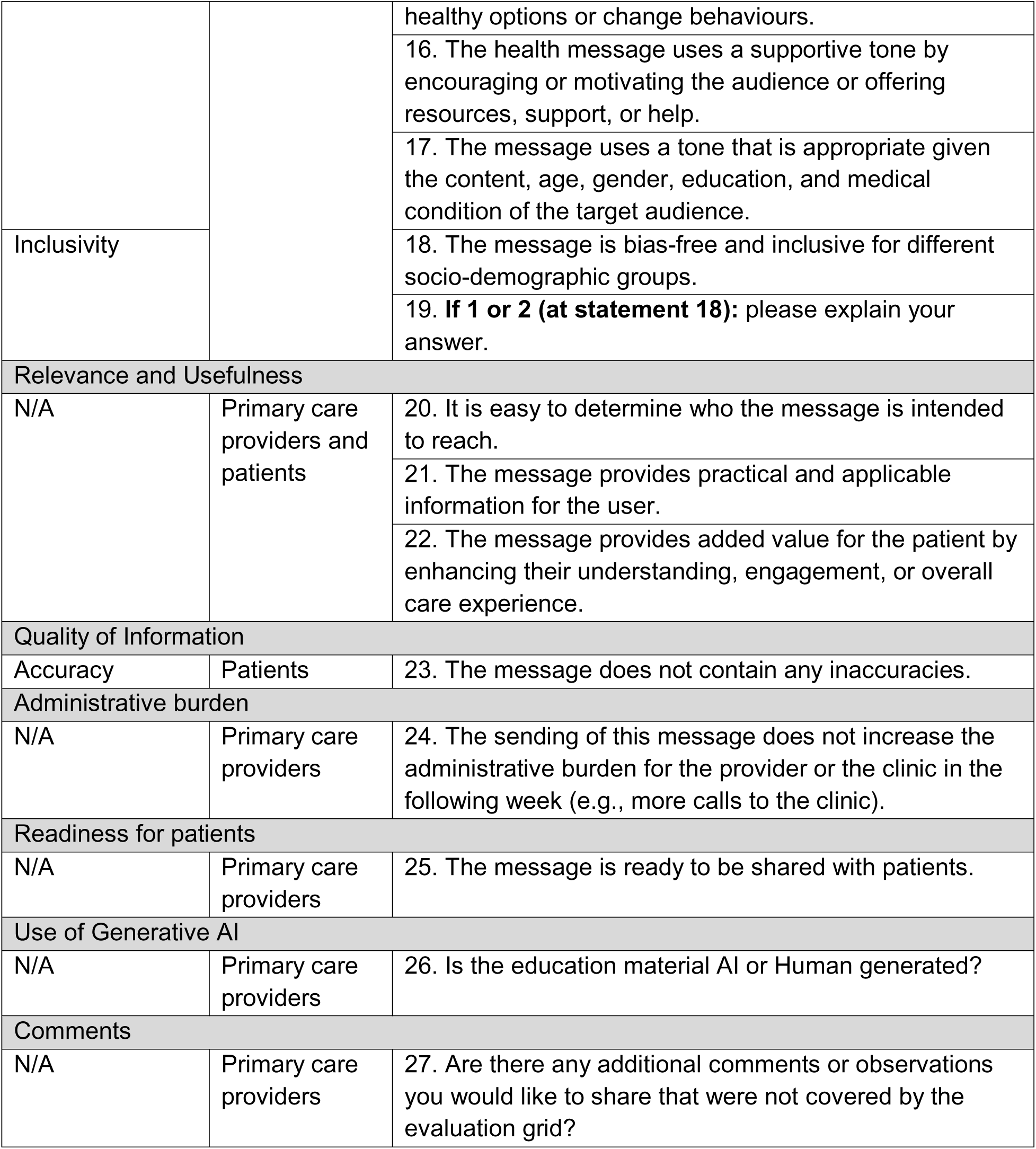
Statements and questions presented in the Evaluation Grid used to assess messages.

At the end of the Evaluation Grid, both primary care providers and patients will be asked to predict whether the message was generated by humans or GenAI (question 26; 0: “AI-generated” and 1: “Human-generated”) and to provide any additional comments they wish to share (question 27; text box).

All evaluations will be completed online using REDCap [80,81] surveys. Each month, the 16 digital health messages followed by their respective Evaluation Grids will be shown in sequential order (see online supplemental materials 2 [English] and 3 [French]). Once evaluations have been submitted for a given message, panelists can no longer go back and modify their answers.

Before data collection, panelists received a group training in French or English on the study objectives and tasks, including how to use the Evaluation Grid. Practice rounds evaluating messages were conducted to ensure all panelists felt comfortable using the Evaluation Grid and had no further questions on its different components. The recordings of those sessions and the materials presented were shared with panelists who were not able to join at the time or joined the study later on, and made available for any follow-up questions.

### Sample size

We performed a simulation to estimate the sample size required for this crossover, noninferiority trial (see online supplemental material 4). We generated synthetic data under the following assumptions: an intercept of the expected understandability score of 6 in the control condition (human-generated messages), a true treatment effect of 0 (consistent with a noninferiority design), a range of -5 to 5 for individual topic understandability, a random effect standard deviation (sd) of 0.5, and a random error sd of 0.3. This results in an approximate mean/median understandability score of 8.5, with a sd of 1.8 and an interquartile range of 7.2 and 9.8 (similar to unpublished CPIN understandability data from March 2024). The methodology used to obtain these data is described in Johnston et al., 2025 [9].

Each simulated dataset was analyzed under the following linear mixed-effects model, where random intercepts were generated for the topic and the individual:

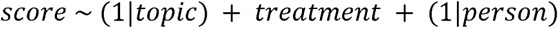

The simulation revealed that with α = 0.05, a total sample size of 40 participants (20 providers and 20 patients) yields >99% of statistical power to conclude noninferiority, assuming a noninferiority margin of 0.5 on the Clarity and Understandability scale. To account for potential attrition and underestimations of our assumptions, we recruited 8 more participants (4 providers and 4 patients).

The simulation was performed using the R (v4.4.3) packages lme4 (v1.1-37) [82] and lmerTest (v3.1-3) [83]. The generated R code can be found on GitHub [84].

### Recruitment of primary care providers and patients

All participants in this study must be able to consent to participate and sign a consent form before enrollment. Individuals with an expertise in primary care, public health, and/or health communication, and who can write in English or French, will be eligible for the Content Team. Primary care providers for the Review Panel must provide community-based care in Canada and be proficient in either written English or French to be eligible. Similarly, eligible primary care patients must be proficient in either written English or French, and be aged 18 years or older. Primary care providers and patients who do not have an email address, a computer, or a cellphone to complete the online REDCap evaluations will be excluded. Furthermore, this study will not include minors or patients who cannot provide informed consent themselves, such as those with advanced dementia.

Primary care providers from various professions (family physicians, registered nurses, nurse practitioners, physiotherapists, pharmacists, etc.) will be recruited from CPIN’s existing network of over 75 contacts, including providers involved in other CPIN studies [8,9]. We aim to recruit at least 20% of primary care providers whose most spoken language at home is French. Primary care providers on the Review Panel will be financially compensated $100 per topic, for a maximum of $4,800 for the entire study. Primary care providers on the Content Team will not be financially compensated for their time.

To recruit primary care patients to the study, we included an additional question in short patient surveys distributed through other CPIN projects [9], briefly explaining the study’s goals and whether patients wished to be contacted for this initiative. Those who accepted to be contacted were sent an email to which they could reply if they were still interested to participate. As the survey answers were linked to EMR-extracted data (age, sex, language in the EMR, three first characters of the postal code, and number of medical conditions), we contacted patients with different sociodemographic characteristics to diversify the study population. As CPIN regularly receives emails from patients who want to learn more about CPIN or to subscribe to CPIN services, we also sent a recruitment email to patients who had reached out to us in the past year. We aim to recruit at least 20% of primary care patients whose most spoken language at home is French. Patients on the Review Panel will be financially compensated $25 per topic, for a maximum of $1,200 for the entire study.

Participants can withdraw at any point during the study. Panelists who withdraw from the study will be financially compensated for the evaluations completed. If necessary, additional primary care providers and patients will be recruited during the study to maintain the targeted sample size in the case of attrition. Panelists who will be recruited after data collection has begun will receive the same training materials and be able to catch-up on previous evaluations to ensure they provide a complete set (Figure 2).

### Randomization and blinding

Each month, panelists will evaluate 4 messages per topic for a total of 16 messages (192 messages and 48 topics over 12 months). Messages will be presented in a sequential, randomly determined order that varies from one individual to another (Figure 1). Randomization is done at the topic level using a block size of 48 to make them unique to each individual, stratified by the type of evaluation (primary care provider and patient). Short and long messages will come from the same generation source (“human” or “AI”), as described in the “Evaluation of digital health messages by provider and patient Review Panels” section.

Before data collection began, the Research Coordinator from CPIN produced two computer-generated permutated-block randomization lists (one for primary care providers and one for patients on the Review Panels). The randomization lists were sent to the CPIN Research Assistant, who is responsible for creating and sending the REDCap evaluation surveys. The CPIN Research Coordinator, other study team members, and the panelists are blinded to the randomization order.

### Outcomes

The primary outcome is the Clarity and Understandability score, which is an important predictor of efficient and effective patient communication [85,86], and will be measured on three Likert scale statements from the study Evaluation Grid (total possible score between 3 to 12). The secondary outcomes include the four message category scores: i) quality of information (scores between 5 to 20 [providers] or 1 to 4 [patients]), ii) adaptability (scores between 7 to 28), iii) relevance and usefulness (scores between 3 to 12), and iv) readiness to be shared with patients (scores between 1 to 4). Other secondary outcomes are each individual criterion score (scores between 1 to 4). Statements that collect qualitative data (#19 and #27) or do not use a Likert scale (#26) will be excluded. Statements on tone identification (#11 to #16) will be excluded, as identifying a tone does not inherently indicate whether the message performed well or poorly. Only the statement assessing the appropriateness of the tone(s) used will contribute to the reported scores.

### Statistical analyses

Statistical analyses will be performed after data collection and quality assurance has been completed. We will report the number and percentage of panelists that were screened, contacted, enrolled, and who withdrew from the study. Continuous variables will be summarized using one or multiple of the following descriptive statistics: n, percentage, mean, standard deviation, median, maximum, and minimum. Number and percentage of observed levels will be reported for all categorial measures. Summary tables will be annotated with the relevant total population size, including any missing observations. Linear mixed-effects models will be used for analyses of the primary and secondary outcomes, accounting for clustering levels at the topic, month, and of the individual.

### Data monitoring and missing data

Quality assurance analyses will be conducted continuously throughout the study to ensure data monitoring. An R script will be used to verify that panelists complete all 16 evaluations for the month, and will flag participants where one or multiple message evaluations have been omitted (i.e., none of the questions have been answered for a given message). The CPIN Research Coordinator will then notify the CPIN Research Assistant, who will go back to the panelist and ask them to provide the missing evaluations. Furthermore, another R script will be used to verify that panelists do not always put the same score or “Not Applicable” for all messages, which may indicate a misunderstanding of the Evaluation Grid. If panelists forgot or omitted to answer one or multiple questions of a given message evaluation, this will be considered as “missing data.”

If a panelist withdraws at any point during the study, the partially completed data will be kept for analyses. Panelists are only considered enrolled once they have signed the consent form, completed the initial baseline survey, and evaluated at least one message.

### Patient and public involvement in research

Patients and/or the public were not involved in the design, conduct, reporting, or dissemination plans of this research.

## ETHICS AND DISSEMINATION

The Research Ethics Board (REB) of the Hôpital Montfort (24-25-11-038) formally approved the study and its documentation (i.e., the protocol, recruitment and measurement tools, and consent forms) in December 2024. The University of Ottawa REB (S-12-24-11153) subsequently granted approval based on the Montfort REB’s decision. Any amendments to the study protocol and/or any of its documentation have been and will continue to be submitted to the Hôpital Montfort REB for formal approval.

Any potential AI-CARE participant (Content Team and Review Panels) must read and sign a consent form before enrollment. As sociodemographic information of primary care providers and patients will be collected, names and initials will be anonymized and replaced by a code (PRX for primary care providers, PAX for patients) in datasets. Only certain members of the research team will have access to the code file list for identification, follow-up, and financial compensation purposes. Data with identifiable information will be kept safely on a secure server at the Hôpital Montfort, while anonymized data without identifiable information will be kept on the Hôpital Montfort SharePoint site. Message scores may be linked with sociodemographic information, although names and initials will be removed during the linkage and replaced by a code. Reported data will be grouped and no identifying information will be used at any point during dissemination. Anonymized data for this study may be used for future research purposes. An REB’s approval will be required for any future studies using these data.

This study represents no risk for the Content Team members or panelists. Messages deemed inaccurate, inappropriate, or not ready to be shared with patients will never be publicly available or be sent out to primary care patients beyond those in the panel. Any message sent to patients requires supervision and approval of their primary care provider. In the consent forms, Review Panels were notified that some messages may contain inaccurate information.

The methodology of this study was presented in March 2025 at the *Journée Recherche Montfort 2025* (local conference) and in April 2025 at the *University of Ottawa’s Department of Family Medicine Retreat*. At the end of the data collection, the research team will publish the study results in peer-reviewed scientific journals, and present them to regional, national, and international conferences. Following data collection, panelists may also receive a list of which messages were generated by humans and AI, and whether they accurately identified the message’s generation source.

### Trial status

The AI-CARE study was registered on May 30, 2025, on CinicalTrials.gov (<u>NCT06997107</u>). Data collection for this study began on May 30, 2025, and is expected to be completed at the end of April 2026. The current protocol version approved by the Hôpital Montfort REB is V7 (July 24, 2025). Amendments to the protocol were minor and included changes to our recruitment approach (addition of recruitment emails) and measurement tools (addition of the AI Usage Survey for primary care provider panelists and translation of the measurement tools from English to French).

### Data availability statement

The R scripts used for the randomization and sample size simulation can be found on GitHub [84]. The datasets generated or analyzed during this study will not be publicly available. Anonymized data may be shared with external researchers upon request and approval by a Research Ethics Board.

## DISCUSSION

This manuscript outlines the study protocol of the AI-CARE study, a randomized controlled trial assessing whether AI-generated health messages are evaluated as noninferior compared to those generated by humans. The results of this study will provide insight into whether GenAI can meet appropriate standards of care for accuracy in primary care and public health content. Additionally, this study will examine whether GenAI can function as a tool to facilitate or extend tailored patient communication in primary care.

To our knowledge, this study will be among the first to evaluate AI- and human-generated health messages using the perspectives of both primary care providers and patients in parallel, recognized as an essential collaboration to improving primary care [87,88]. Our study population is diverse, with primary care providers on the Content Team and the Review Panel being recruited from different professions and provinces in Canada, and patients on the Review Panel representing a range of sociodemographic characteristics, including age, sex, language, and rurality.

This is also one of the first studies that will thoroughly evaluate the human- and AI-generated messages on such a breadth and depth of communication elements: i) quality of information (specifically accuracy, reliability, and completeness), ii) adaptation the target audience (broken down as clarity and understandability, appropriate emotional and rational appeal, tone, and inclusivity), iii) relevance and usefulness (or “actionability”), and iv) readiness to be shared with patients. AI-CARE was not designed to generate responsive or individualized content, such as in crafting responses to patient inbox messages [24]. Instead, it will investigate GenAI capacities of crafting proactive health information on a wide variety of topics and targeting different patient populations, which few studies have done to date.

Cognitive fatigue may be observed in some participants and could affect the accuracy of their evaluations. However, rather than requiring panelists to review all 192 messages in a short period to ensure content diversity, we reduced the workload and fatigue by having them evaluate 16 messages per month over one year (approximately 1 to 4 hours of work monthly). This allowed us to include primary care providers currently practicing in Canada, who may not have had the time to participate in the study if the burden had been too high, and to cover time-sensitive and non-time-sensitive topics, replicating a one-year cycle of multiple seasons. The within-participant experimental design of this research also limits variability due to individual differences and subjectivity in the understanding of the Evaluation Grid, and reduces the sample size required to achieve sufficient statistical power.

We acknowledge some potential limitations to this study design. For instance, although prompts are standardized, AI-generated messages will be crafted using only one LLM (ChatGPT), which risks introducing LLM-specific bias [89,90]. Moreover, as models are rapidly evolving within a same LLM, studies have shown there is inter-model variability [64–66,91,92]. In August 2025, OpenAI described GPT-5 as “their best model yet for health-related questions,” and indicated improvement in writing style to provide more adapted, reliable, and precise answers [93]. Some heterogeneity may thus be expected between AI-generated messages across topics.

Nevertheless, although GenAI’s performance may improve as the study progresses and be linked to the model used, human experts may similarly improve, as both will benefit from accumulated experience over time. This observation would be reflected in the collected data. Bias is mitigated by our prompt engineering approaches, such as asking ChatGPT to generate three different versions of the message and improving on its previous outputs, using the same prompts across models, generating messages in different threads to avoid contexts being mixed up, and giving human experts and the GenAI tool the same training materials and instructions.

Due to our recruitment strategy, there are also risks of introducing selection bias, as these participants are already research-oriented and interested in AI and/or digital tools. Consequently, this could affect, positively or negatively, the evaluations of human- and AI-generated messages. However, this approach is acceptable considering that this is an exploratory protocol and that messages will not be sent out to patients in a context of care outside of the patient panelists. Moreover, because recruitment for patient and primary care provider panelists was focused in Ontario, the racial diversity of our study population may be limited.

## ETHICS STATEMENTS

### Patient consent for publication

Not applicable.

## ACKNOWLEDGEMENTS

The authors would like to thank Stéphanie Chenail, Laurie Woolley, Miryam Duquet, and Mathieu Côté from the Institut du Savoir Montfort. They would also like to thank Kevin Haaland from Cliniconex and Richard Webster from the CHEO Research Institute for their input in the trial design, Nick Barrowman from the CHEO Research Institute for his help with the sample size simulation, and Chantal Sauvé from the Eastern Ontario Health Unit for the translation of messages.

## AUTHOR CONTRIBUTIONS

AL coordinates the project, designed the trial and the analysis plan, and wrote the original draft of the manuscript. SAK advised on the trial design and developed the analysis plan. BRGT helped coordinate the project, design the trial, and write the original draft of the manuscript. GS helped design the trial. JT contributed to the trial design and recruitment methods. DA contributed to the trial design. SG and WH conceived the study and helped design the trial and acquire funding. SJ conceived the study, designed the trial, contributed to the analysis plan and the writing of the original draft of the manuscript, and acquired funding. All authors have read and approved the final version of the manuscript.

OpenAI ChatGPT was used sporadically to improve readability and correct grammatical errors of this manuscript. The generated output was reviewed, edited, and validated by the authors.

