## Supplementary material for "A double-blind, crossover, non-inferiority randomized controlled trial where primary care providers and patients compare human- and AI-generated digital health messages: the AI-CARE study protocol": Online supplemental material 1

**Supplemental material 1. Survey questions.**

* Identify questions where answers are required

**Initial survey (sociodemographic information collected at onboarding)**

1. **In what language do you want to complete the survey** (*will change the language of the survey automatically*)**?***

- English
- Français

2. **What is your last name?* _______________**
3. **What are your initials?* _______________**
4. **Are you a provider or a patient/caregiver?***

- Provider
- Patient/caregiver

5. **What language do you speak most often at home?** *(Indicate more than one language only if they are spoken equally at home)*

- English
- French
- Other

< If **Other** >

5.1 **Please specify: _______________**
5.2 **In what official language of Canada are you most comfortable receiving your healthcare services?**
- English
- French

6. **People in Canada come from many racial or cultural groups. You may belong to more than one group on the following list. Please select all that apply:**

- Middle Eastern or North African (Lebanese, Algerian, Iraqi, Syrian, etc.)
- Black (African Canadian, Caribbean, African origin, etc.)
- East Asian (China, Hong Kong, Japan, North Korea, South Korea, etc.)
- Indigenous (First Nations, Métis or Inuit)
- Latin American (Mexico, Haiti, Uruguay, etc.)
- South Asian (East Indian, Sri Lankan, etc.)
- South-East Asian (Vietnamese, Cambodian, etc.)
- West Asian (Iranian, Afghan, etc.)
- White/Caucasian
- Prefer not to say
- Or another group? Please specify: _______________

7. **We want to better understand our participants and ensure our work is inclusive. Which of the following best describes your sexual orientation?**

- Heterosexual (Straight)
- Lesbian
- Gay
- Bisexual
- Pansexual
- Asexual
- Queer
- Questioning or unsure
- Prefer not to say
- Other? Please specify: _______________

< Questions 8 will be asked if they selected **Patient/caregiver** >

8. **What is your highest level of education?**

- Grade 12 or less
- I went to college or university but didn’t get my degree
- I have a college or university degree
- I have an advanced professional or postgraduate degree (Masters, PhD, physician, lawyer, etc.)
- Other? Please specify: _______________

9. **What is your gender?**

- Man
- Woman
- Non-binary
- Other
- Prefer not to say

10. **How old are you? _______________**
11. **What are the first three digits of your postal code? _______________**

< Questions 12 will be asked if they selected **Patient/caregiver** >

12. **What is your gross annual income?**

- $0 to $24,999
- $25,000 to $49,999
- $50,000 to $74,999
- $75,000 to $99,999
- $100,000 to $124,999
- $125,000 to $149,999
- $150,000+

< Questions 13 to 16 will be asked if they selected **Provider** >

13. **What is your profession?**

- Family doctor
- Nurse practitioner
- Registered nurse
- Pharmacist
- Social worker
- Psychologist
- Dietician or Nutritionist
- Physiotherapist
- Other professional qualifications? Please specify: _______________

14. **What are the first three digits of the postal code of your workplace? _______________**
15. **How many years have you been practicing in your current field?**

- Less than 1 year
- 1 to 5 years
- 6 to 10 years
- 11 to 15 years
- 16 years to 20 years
- More than 20 years

< Questions 16 will be asked if they selected **Patient/caregiver** >

16. **Do you identify as a caregiver for someone due to their health needs, disability, aging, or other reasons?**

- Yes, I am a primary caregiver (responsible for most of the care).
- Yes, I am a secondary caregiver (I help but am not the main provider of care).
- No, I do not provide care for anyone.

**AI Usage of Primary Care Providers of the Review Panels**

1. How do you currently use generate AI in your clinical practice? (e.g., drafting notes, patient education, summarizing records)
2. Do you currently use generative AI to help create or adapt patient education materials? If yes, how?
3. **If you answered ‘Yes’ at question 2:** Roughly how much time do you usually have to prepare or review these materials for each patient? (e.g., 2 minutes? 10 minutes? 30 minutes? 1 hour?)
4. **If you answered ‘Yes’ at question 2:** When using AI, do you prefer giving it detailed, step-by-step instructions – or using a pre-set template or standard prompt? Why?

*(You can indicate ‘N/A’ if you don’t know the difference between both approaches)*
