## Supplementary material for "A double-blind, crossover, non-inferiority randomized controlled trial where primary care providers and patients compare human- and AI-generated digital health messages: the AI-CARE study protocol": Online supplemental material 2

**Supplemental material 2. Example of a REDCap survey.**

* Identify questions where answers are required

1. **In what language do you want to complete the survey** (*will change the language of the survey automatically*)**?***

- English
- Français

2. **What is your last name?* _______________**
3. **What are your initials?* _______________**
4. **Are you part of the providers’ or patients’ review panel?***

- Providers’
- Patients’

Page break

**Topic 1: short message 1 (AI or human-generated, randomly assigned)**

*Message here*

**Evaluation grid:**

|  | | **Strongly disagree**  **1** | **Disagree**  **2** | **Agree**  **3** | **Strongly agree**  **4** | **Not Applicable** |
| --- | --- | --- | --- | --- | --- | --- |
| **Quality of Information (providers)** | | | | | | |
| < If **provider** was selected> | | | | | | |
| Accuracy | The information presented in this message is in factual agreement with the evidence-based reference. |  |  |  |  |  |
|  | No unnecessary or inappropriate information was added to the message. |  |  |  |  |  |
| Reliability | The references cited are reliable and timely. |  |  |  |  |  |
|  | The information provided is consistent with current scientific knowledge. |  |  |  |  |  |
| Completeness | Given its length, no necessary information was omitted from the message. |  |  |  |  |  |
| **Adaptability** | | | | | | |
| Clarity and understandability | The content of the message is appropriate given the age, gender, education, and medical condition of the target audience. |  |  |  |  |  |
|  | The message is structured logically and coherently. |  |  |  |  |  |
|  | The message does not contain unnecessary technical jargon. |  |  |  |  |  |
| Appropriate emotional appeal | The message uses **the appropriate emotional appeal for the target audience** (positive, negative, affiliation, humour) and positive benefits (security, pride, joy, fun, positive mood) to persuade people to adopt behaviours that favour social and individual well-being. |  |  |  |  |  |
| Appropriate Rational appeal | The message uses **the appropriate rational appeal for the target audience** using factual arguments by making a direct presentation of the facts to appeal to reason, thinking and awareness. |  |  |  |  |  |
| Tone | The health message uses an ***empathetic tone*** which is conveyed as comprehensive, reassuring, and helpful. |  |  |  |  |  |
|  | The health message uses an ***engaging tone*** to develop affinity with the subject. |  |  |  |  |  |
|  | The health message uses a ***directive tone*** by telling what to do and what not to do, using educational reasoning. |  |  |  |  |  |
|  | The health message uses a ***solution-based tone*** providing tips and ideas to encourage the target audience to do something, by showing them how. |  |  |  |  |  |
|  | The health message uses a ***substitution-based tone*** providing tips and ideas or showing how to choose healthy options or change behaviours. |  |  |  |  |  |
|  | The health message uses a ***supportive tone*** by encouraging or motivating the audience or offering resources, support, or help. |  |  |  |  |  |
|  | The message uses a tone that is appropriate given the content, age, gender, education, and medical condition of the target audience. |  |  |  |  |  |
| Inclusivity | The message is bias-free and inclusive for different socio-demographic groups. |  |  |  |  |  |
|  | If **1 or 2** (for the previous question): please explain your answer. |  | | | | |
| **Relevance and Usefulness** | | | | | | |
|  | It is easy to determine who the message is intended to reach. |  |  |  |  |  |
|  | The message provides practical and applicable information for the user. |  |  |  |  |  |
|  | The message provides added value for the patient by enhancing their understanding, engagement, or overall care experience. |  |  |  |  |  |
| **Quality of Information (patients)** | | | | | | |
| < If **patient** was selected > | | | | | | |
| Accuracy (for patients only; at the end) | The message does not contain any inaccuracies. |  |  |  |  |  |
| **Administrative burden (providers)** | | | | | | |
| < If **provider** was selected > | | | | | | |
|  | The sending of this message does not increase the administrative burden for the provider or the clinic in the following week (e.g., more calls to the clinic). |  |  |  |  | **Quality of Information (patients)** |
| **Readiness for Patients** | | | | | | |
|  | The message is ready to be shared with patients. |  |  |  |  |  |

|  |  | **AI generated**  **0** | **Human generated**  **1** |
| --- | --- | --- | --- |
| **Use of Generative AI** | | | |
|  | Is the education material AI or Human generated? |  |  |

**Comments**

Are there any additional comments or observations you would like to share that were not covered by the evaluation grid?

Page break

**Topic 1: long message 1 (AI or human-generated, randomly assigned)**

*Message here*

Evaluation Grid

Page break

**Topic 2: short message 1 (AI or human-generated, randomly assigned)**

*Message here*

Evaluation Grid

Page break

**Topic 2: long message 1 (AI or human-generated, randomly assigned)**

*Message here*

Evaluation Grid

Page break

**Topic 3: short message 1 (AI or human-generated, randomly assigned)**

*Message here*

Evaluation Grid

Page break

**Topic 3: long message 1 (AI or human-generated, randomly assigned)**

*Message here*

Evaluation Grid

Page break

**Topic 4: short message 1 (AI or human-generated, randomly assigned)**

*Message here*

Evaluation Grid

Page break

**Topic 4: long message 1 (AI or human-generated, randomly assigned)**

*Message here*

Evaluation Grid

**Topic 1: short message 2 (AI or human-generated, alternate of Topic 1 – message 1 randomization)**

*Message here*

Evaluation Grid

Page break

**Topic 1: long message 2 (AI or human-generated, alternate of Topic 1 – message 1 randomization)**

*Message here*

Evaluation Grid

Page break

**Topic 2: short message 2 (AI or human-generated, alternate of Topic 2 – message 1 randomization)**

*Message here*

Evaluation Grid

Page break

**Topic 2: long message 2 (AI or human-generated, alternate of Topic 2 – message 1 randomization)**

*Message here*

Evaluation Grid

Page break

**Topic 3: short message 2 (AI or human-generated, alternate of Topic 3 – message 1 randomization)**

*Message here*

Evaluation Grid

Page break

**Topic 3: long message 2 (AI or human-generated, alternate of Topic 3 – message 1 randomization)**

*Message here*

Evaluation Grid

Page break

**Topic 4: short message 2 (AI or human-generated, alternate of Topic 4 – message 1 randomization)**

*Message here*

Evaluation Grid

Page break

**Topic 4: long message 2 (AI or human-generated, alternate of Topic 4 – message 1 randomization)**

*Message here*

Evaluation Grid
