## Supplementary material for "A double-blind, crossover, non-inferiority randomized controlled trial where primary care providers and patients compare human- and AI-generated digital health messages: the AI-CARE study protocol": Online supplemental material 4

Sample size simulation

2025-12-15

### Load libraries

library(lme4)

library(lmerTest)

### Modify values

```
# Modify values
G <- 1000 # Number of iterations
n_people <- 40
n_topics <- 48

re_sd <- 0.5
error_sd <- 0.3
intercept <- 6   # Expected score in the control group (human-generated messages)
treatment_effect <- 0 # Consistent with a noninferiority trial design
```

### Create a function to simulate the data of one person

```
get_newperson <- function(person_number,intercept,topic_df,
  treatment_effect,re_sd,error_sd) {

  #browser()
  n_topics <- length(topic_understandability)

  tu <- c(topic_df$topic_understandability,topic_df$topic_understandability)
  treatment <- c(rep(0,n_topics),rep(1,n_topics))

    re <- rnorm(1,mean=0,sd=re_sd)
    error <- rnorm(2*n_topics,mean=0,sd=error_sd)
    score_vector <- intercept + re + tu + treatment*treatment_effect + error
    data.frame(person=rep(person_number,2*n_topics),
      treatment=treatment,topic=c(topic_df$topic,topic_df$topic), score=score_vector)
}
```

### Create a function to simulate the entire dataset

```
# Create a function to simulate data
generateRealization <- function(n_people,intercept,topic_df,treatment_effect,re_sd,error_sd) {
  fulldata <- NULL
  for (i in 1:n_people) {
  person <- get_newperson(i,intercept=intercept,
    topic_df = topic_df, treatment_effect=treatment_effect,
    re_sd=re_sd,error_sd=error_sd)
  #browser()
  fulldata <- rbind(fulldata,person)
  }
  fulldata
}
```

### Analyzed the simulated dataset under the following model

```
topic_names <- paste0("topic",1:n_topics)

topic <- factor(topic_names,levels=topic_names)

topic_understandability <- seq(0,5,length=n_topics)

topic_df <- data.frame(topic, topic_understandability)

fulldata <- generateRealization(n_people=n_people,intercept=intercept,
  topic_df=topic_df,treatment_effect=treatment_effect,
  re_sd=re_sd,error_sd=error_sd)

overallMean <- mean(fulldata$score)

fit <- lmer(score ~ (1|topic) + treatment + (1|person), data=fulldata)
fit
## Linear mixed model fit by REML ['lmerModLmerTest']
## Formula: score ~ (1 | topic) + treatment + (1 | person)
##    Data: fulldata
## REML criterion at convergence: 2197.315
## Random effects:
##  Groups   Name        Std.Dev.
##  topic    (Intercept) 1.4842
##  person   (Intercept) 0.5239
##  Residual             0.2982
## Number of obs: 3840, groups:  topic, 48; person, 40
## Fixed Effects:
## (Intercept)    treatment
##   8.4638107   -0.0000944
ci <- confint(fit)
## Computing profile confidence intervals ...
ci
##                   2.5 %     97.5 %
## .sig01       1.21896040 1.82236279
## .sig02       0.42524661 0.66446039
## .sigma       0.29151607 0.30500954
## (Intercept)  8.01003643 8.91758504
## treatment   -0.01895856 0.01876976
```

### Analyze G = 1,000 iterations

```
set.seed(1234)
lower <- upper <- rep(NA, G)

for (i in 1:G) {
  fulldata <- generateRealization(
    n_people = n_people, intercept = intercept,
    topic_df = topic_df, treatment_effect = treatment_effect,
    re_sd = re_sd, error_sd + error_sd
  )

  fit <- tryCatch(
    lmer(score ~ (1 | topic) + treatment + (1 | person),
      data = fulldata,
      control = lmerControl(
        optimizer = "bobyqa", optCtrl = list(
          maxfun = 2e5
        )
      )
    ),
    error = function(msg) {
      message(msg)
      NULL
    }
  )

  if (is.null(fit)) {
    lower[i] <- NA
    upper[i] <- NA
  } else {
    ci <- confint(fit)

    lower[i] <- ci["treatment", "2.5 %"]
    upper[i] <- ci["treatment", "97.5 %"]
  }
}
```

### Compute power assuming a 0.5 noninferiority margin

```
delta <- -0.5

# Compute power
power <- length(which(lower > -0.5))/length(lower)
print(paste0("Computed power was: ", power))
## [1] "Computed power was: 1"
# Save data
save.image("251215_Simulations.RData")
```
